# Targeted and optimized multi-channel transcranial direct current stimulation for focal epilepsy: An N-of-1 trial

**DOI:** 10.1101/2023.09.05.23295060

**Authors:** Marios Antonakakis, Fabian Kaiser, Stefan Rampp, Stjepana Kovac, Heinz Wiendl, Walter Stummer, Joachim Gross, Christoph Kellinghaus, Maryam Khaleghi-Ghadiri, Gabriel Möddel, Carsten H. Wolters

## Abstract

Transcranial direct current stimulation (tDCS) has been used to noninvasively reduce epileptic activity in focal epilepsy. In this proof-of-principle N-of-1 trial in a patient with drug-resistant focal epilepsy, we propose distributed constrained maximum intensity (D-CMI) for individually targeted and optimized multi-channel (mc-) tDCS to reduce epileptic activity. Combined electro- and magnetoencephalography (EMEG) source analysis in a realistic calibrated head model defines location and orientation of the target epileptogenic source. Converging evidence for this determination is achieved by retrospective identification of a cortical malformation in magnetic resonance imaging and by successful EMEG-guided invasive EEG. We applied D-CMI in a double-blind, sham-controlled stimulation experiment. In two stimulation weeks, either D-CMI or sham stimulation with 4 mA injection current were applied twice every week-day for 20 min each, with a 20 min pause in between. EEG was recorded 1 h before and after stimulation. For D-CMI, we find a highly significant reduction in IED frequency (p < 0.0001) marked by three experts of on average 37% to 81% over the five days of stimulation (mean ± SD: 58% ± 19%), while this is not the case for sham. The proposed procedure was well-tolerated and parameterizes a group clinical trial (Study registration number: DRKS00029384).

## 1. Introduction

Worldwide, around fifty million people suffer from epilepsy, making it one of the most common neurological diseases globally (WHO, 2022). The majority of them are affected by focal epilepsy (Beghi, 2020). If focal epilepsy does not respond to two or more adequate anti-seizure medications it is considered as refractory (Kwan et al., 2010). The focus of this work is the non-invasive treatment of refractory focal epilepsy by means of transcranial direct current stimulation (tDCS) (Antal et al., 2017; Boon et al., 2018; Yang et al., 2020; Kaufmann et al., 2021).

tDCS is based on a sufficiently accurate localization of the target *epileptogenic zone*, defined as the area of cortex indispensable for the generation of clinical seizures (Rosenow and Lüders, 2001). tDCS is usually applied via standard bipolar stimulation, i.e., two large patch electrodes, one cathode and one anode, but controversial effects were reported in drug-resistant epilepsy (Boon et al., 2018). However, using cathodal tDCS, i.e., tDCS montages with the cathode over the target, reductions of epileptic activity and seizure frequency or duration have already been reported in preliminary studies (Kaye et al., 2023; Ashrafzadeh et al., 2023; Rezakhani et al., 2022; Kaufmann et al., 2021; Yang et al., 2020; Holmes et al., 2019; San-Juan et al., 2017).

Compared to standard bipolar or cathodal tDCS, stimulation may be more effective if applied via a multi-channel tDCS (mc-tDCS) cap, with a custom-tailored distribution of stimulation electrodes and individualized electric field modeling (Kaye et al., 2023; Kasten et al., 2019; Liu et al., 2018; Ruffini et al., 2014; Sadleir et al., 2012). mc-tDCS injected currents should not only lead to maximal intensity in the target region (IT), but currents should additionally have maximal directionality (DIR), i.e., be oriented parallel (anti-parallel for inhibition in epilepsy) to the target vector orientation (Evans et al., 2022; Zulkifly et al., 2022; Seo et al., 2017; Krieg et al., 2015). A further goal should be to minimize intensity in non-target regions (INT) and thereby possible side effects of the stimulation (focality).

In this *proof-of-concept N-of-1* approach, we use our recently developed distributed constrained maximum intensity (D-CMI) mc-tDCS approach (Khan et al., 2022, 2023) for the first time for individually targeted and optimized neurostimulation in a refractory focal epilepsy patient with the goals to non-invasively reduce epileptic activity and to parameterize our processing pipelines as a preparation for a future study on a group of focal epilepsy patients. Our computer simulation (Khan et al., 2022) and experimental group study (Khan et al., 2023) showed that D-CMI leads to high IT and DIR, coupled with a relatively low INT, outperforms standard bipolar and cathodal stimulation and leads to better control over stimulation outcomes.

D-CMI requires in a first step an accurate reconstruction of the epileptogenic zone with regard to location and orientation (targeting) and in a second step the calculation of the individual electrode montage for an efficient inhibition of this target zone. For the first step, as there is no way to detect the epileptogenic zone directly, several surrogates must be assessed: Ictal EEG(/MEG) can be used to localize the *seizure onset zone* and interictal EEG/MEG to localize the *irritative zone* (Rosenow and Lüders, 2001). However, (Karoly et al., 2016) showed that interictal epileptiform discharges (IEDs) and seizures demonstrate similar probability distributions suggesting they are not wholly independent processes. Furthermore, (Diamond et al., 2023) showed that IEDs are travelling waves arising from an epileptogenic source and in (Heers et al., 2023) the most frequent IED types were found to be significantly concordant with the seizure onset zone. Therefore, our study will focus on EEG and MEG source analysis of IEDs and we will combine both modalities as they complement each other in terms of their sensitivity profiles (Dassios et al., 2007; Piastra et al., 2021): EEG is more sensitive for the detection of both superficial and deep radial sources (e.g., gyral crowns and sulcal valleys), whereas MEG is more sensitive for detecting tangential sources (e.g., sulcal walls) (Knake et al., 2006; Aydin et al., 2015,2017; Ebersole and Wagner, 2018; Plummer et al., 2019; Duez et al., 2019). It has also been shown that early time points such as the middle of the rising flank of the spike (Lantz et al., 2003) or spike onset (Aydin et al., 2015) must be analyzed in order to not just localize propagation. All these results prompted us in the study at hand to use combined EEG/MEG (EMEG) source analysis of IEDs at spike onset for targeting.

In EMEG source analysis, mathematical tools are used to deduce the localization and orientation of intracranial sources from a magnetic field and/or electrical potential distribution detected at the head surface. Due to the focal onset, we use the cortically-constrained deviation scan (CCDS), a straightforward and robust inverse method (Rullmann et al., 2009; Fuchs et al., 1998). We follow the sub-averaging approach of (Aydin et al., 2015) and compute the center of gravity (*centroid*) from sub-averaged IEDs at spike-onset. We show that our approach will enable accurate targeting regarding both centroid location and orientation. For EEG and MEG forward modeling, we use the Finite Element Method (FEM), which provides high flexibility for simulating the electromagnetic field propagation in our patients’ head model. Our modeling will include the presence of complex tissue geometries, conductivity inhomogeneities as well as skull defects (Benar & Gotman, 2002; Lau et al., 2016). An additional calibration of skull conductivity in our patient’s head model enables the combined analysis of EEG and MEG (Antonakakis et al, 2020). White matter anisotropy (Rullmann et al., 2009; Güllmar et al., 2010) is also modeled, as it particularly affects reconstructed source orientations (Güllmar et al., 2010; Aydin et al., 2015). Source orientation is crucial for our case study, not only because the orientation also contains important localizational information (Salayev et al., 2006), but especially because the mc-tDCS targeting is sensitive to the orientation.

In the following, we present a patient with refractory focal epilepsy in left frontal lobe (Section 2.2). Our first goal is to contribute to the identification of the epileptogenic zone by means of EMEG source analysis of sub-averaged IEDs, guiding the retrospective reinspection of 3D-Flair-MRI as well as invasive EEG for the detection of a cortical malformation close to Broca’s area (Sections 2.3-2.6 and 3.1). Second, we compute an individually D-CMI optimized mc-tDCS montage targeting the centroid of the EMEG reconstructed IEDs and show that it leads to high IT and DIR and relatively low INT (Sections 2.7 and 3.2). Third, we perform a double-blind sham-controlled N-of-1 trial EEG/mc-tDCS/EEG stimulation experiment for our patient (Sections 2.8 and 2.9) with the goal to non-invasively reduce epileptic activity (Section 3.3). The main goal is thus to investigate the effect of our non-invasive personalized transcranial electric stimulation on IED frequency. Our hypothesis is that the real stimulation using the D-CMI montage inhibits the irritative zone and thus reduces the frequency of IEDs in EEG’s measured after stimulation when compared to those measured before, while sham stimulation does not. Finally, we present the positive outcome of our personalized mc-tDCS therapy (Section 3.4), discuss the results in the context of the literature (Section 4) and draw conclusions (Section 5).

## 2. Patient and Methods

### 2.1. Ethics Statement

The study was conducted according to the guidelines of the Declaration of Helsinki and approved by the institution’s ethical review boards (Ethik Kommission der Ärztekammer Westfalen-Lippe und der Westfälischen Wilhelms-Universität Münster, 25.05.2021, Ref. No. 2021-290-f-S; Ethik-Kommission der Friedrich-Alexander-Universität Erlangen-Nürnberg, 20.02.2018, Ref. No. 4453B). The patient gave written informed consent for her data to be used in anonymized form for scientific publications.

### 2.2. Patient

We report on a female patient (age range 25-30) with normal intellectual state and otherwise normal neurological status, who was treated for her epilepsy at the University Hospital Münster between 2018 and 2023. The patient was suffering from focal epilepsy since her adolescence, with a seizure semiology described as disturbed thinking and inability to speak or follow a conversation, with no motor symptoms and no impairment of awareness. The frequency of the seizures was between once per week up to four times per day. No seizures with loss of awareness or motor symptoms, and no tonic-clonic seizures (secondary generalization) were noticed. The patient did not become seizure free despite treatment with multiple anti-seizure medications (levetiracetam, lamotrigine, lacosamide, zonisamide, perampanel, oxcarbazepine). As the criteria were met for pharmacoresistance, a first presurgical evaluation for possible resective epilepsy surgery was performed in 2018 (Figure 1).

**Figure 1.**
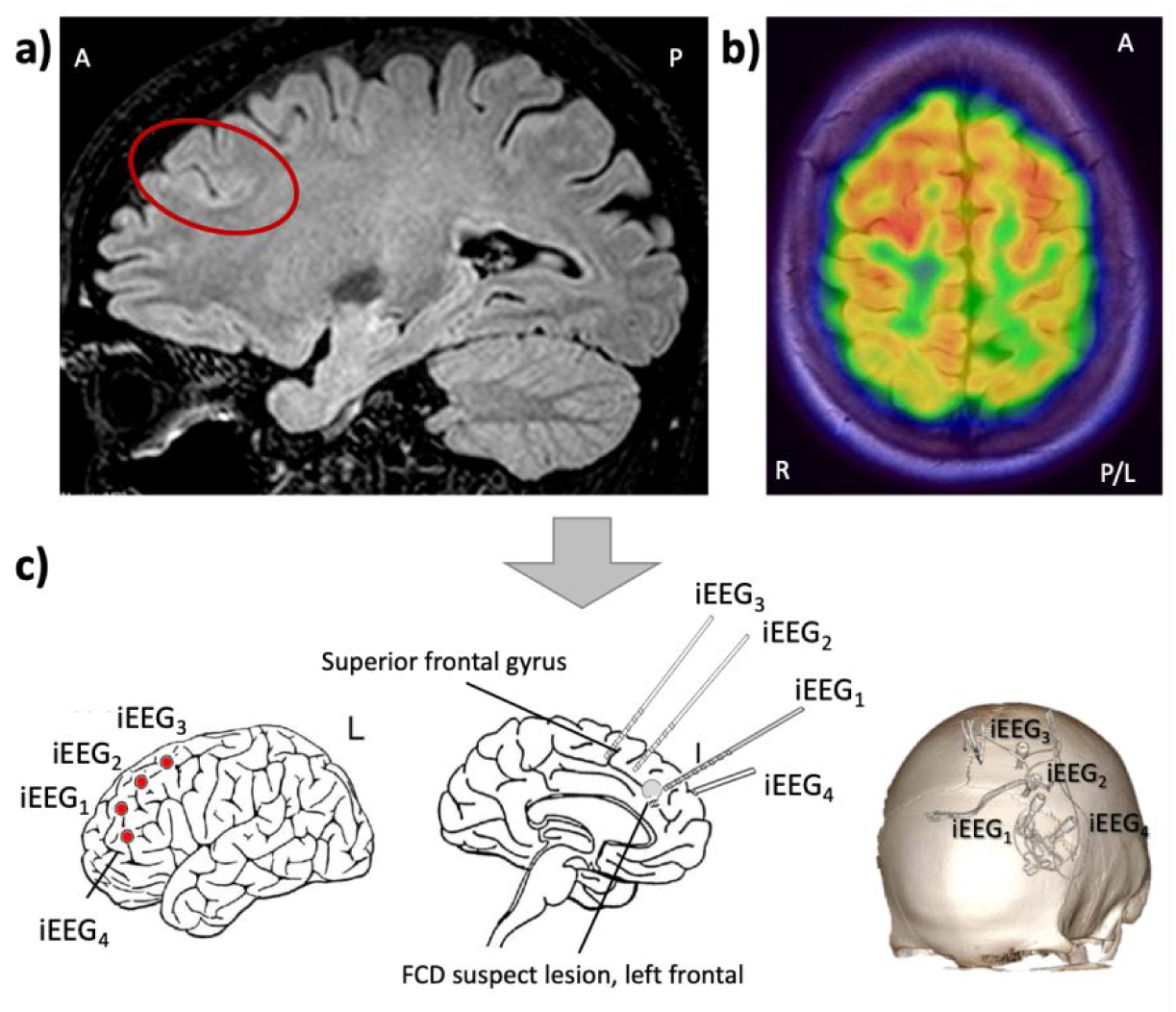
Results of non-invasive presurgical workup and planning of the invasive EEG study in 2018: **a)** a sagittal view of the 3D-FLAIR MRI which was reported as mainly normal, but with a suspicious, unusually deep sulcus in the middle frontal gyrus (indicated by the red circle); **b)** Fluorodeoxyglucose positron emission tomography (FDG-PET) was interpreted as normal, but a very subtle hypometabolism in the left superior frontal gyrus was discussed, suggesting a possible functional deficit zone; **c)** depth electrode placement for the invasive video EEG study (electrodes named iEEG_1_, iEEG_2_, iEEG_3_ and iEEG_4_). Electrodes were implanted into the suspicious deep sulcus and the left superior frontal gyrus. A: anterior, P: posterior, R: right, L: left.

During the initial non-invasive video-EEG monitoring with surface electrodes (10-20 system with additional anterior-temporal and basal-temporal electrodes), electrographic seizure patterns (3 to 5 per day) were recorded in the left frontal area (F3 maximum). The 3D-FLAIR MRI showed a suspicious, unusually deep sulcus in the middle frontal gyrus (Figure 1a). Voxel-based morphometry (VBM) (Huppertz et al., 2005) did not provide clear evidence of a cortical malformation. Fluorodeoxyglucose positron emission tomography (FDG-PET) was reported as normal, but a very subtle hypometabolism in the left superior frontal gyrus was discussed (Figure 1b). Neuropsychological testing was normal. To further improve our hypothesis regarding localization of the epileptogenic zone and to guide optimal electrode placement for an invasive EEG evaluation (Sutherling et al., 2008), a first combined EEG/MEG recording was done (40 minutes recording). The acquisition protocol was the same as described in Section 2.3 below. Unfortunately, no IEDs or seizure patterns could be detected. Thus, the implantation of the invasive EEG electrodes was planned based on the evidence derived from non-invasive video-EEG and MRI alone to confirm the hypothesis of left frontal lobe seizure onset. The implantation of depth brain electrodes was performed in general anesthesia using a stereotactic frame (Leksell Frame G, Elekta, Stockholm, Sweden). Four invasive electrodes (iEEG_1_, iEEG_2_, iEEG_3_ and iEEG_4,_ Figure 1c) were implanted into the suspicious deep sulcus and the left superior frontal gyrus. For accurate visualization of the depth electrode positions (see Figure 1c), cranial Computed Tomography (CT, Siemens SOMATOM Definition, Erlangen, Germany) was acquired. Eight electrographic seizures were recorded, with rhythmic alpha activity in one contact of the depth electrode implanted into the suspicious sulcus (iEEG_4_, contact 5), evolving into rhythmic theta, and spreading to the neighboring electrode contacts. However, IEDs were very infrequent (maximum at iEEG_4_, contact 5). Active contacts were dominated by rhythmic alpha activity and not by low-amplitude fast activity, suggesting that the epileptogenic zone was not captured, and that the recorded seizure patterns represented only spread patterns. Hence, the invasive evaluation was not conclusive. Most likely, the seizure onset zone was very close, but with some distance to contact 5 of electrode iEEG_4_. Therefore, the depth electrodes were explanted without resection. Optimization of medical therapy was recommended. After some unsuccessful drug trials, the patient was not seizure-free a year later, in 2019. We then scheduled a second combined EEG/MEG recording. This time, to increase the probability of documenting epileptiform discharges, anti-seizure medication was tapered 3 days before the recording, and the patient was sleep-deprived. During the recording, over one thousand interictal spikes with maximum in the left frontal region were observed within 40 minutes.

### 2.3. Electrophysiological Measurements and IED Detection

We acquired simultaneous EEG and MEG recordings from the patient in a magnetically shielded room. Eighty-one AgCl sintered ring electrodes (EASYCAP GmbH, Herrsching, Germany, 74 channel EEG plus additional 7 Electro-oculogramm and -cardiogram channels to detect eye movements and heart artifacts) were conducted for EEG and a whole-head MEG system was used (OMEGA2005, CTF, VSM MedTech Ltd., Canada) with 275 axial gradiometers and 29 reference coils (for active noise cancellation using third order synthetic gradiometers). Before measurements, the electrode positions of the EEG cap were digitized using a Polhemus device (FASTRAK, Polhemus Incorporated, Colchester, Vermont, U.S.A.). All EEG/MEG and MRI measurements were done in supine position to reduce head movements and prevent erroneous cerebrospinal fluid (CSF) effects due to brain shift when combining EEG/MEG and MRI (Rice et al., 2013).

Six runs were acquired in total. The first run was 7 minutes long measuring somatosensory evoked potentials and fields (SEP and SEF) for head model calibration with regard to individual skull conductivity (Antonakakis et al., 2020). In the remaining five runs spontaneous activity was acquired with the goal to measure epileptiform activity. The EEG/MEG measurement protocol and the preprocessing steps for reducing the non-cerebral activity was the same as described in (Aydin et al., 2017).

Epileptiform activity (spikes) was manually identified and marked by two experienced epileptologists (authors GM and SR) on the artifact-reduced EEG and MEG continuous data. For the EEG modality, a referential montage (common average) was used. One thousand fifty spikes were selected for further analysis, choosing the common spikes that both evaluators marked. The individual spikes were aligned around zero (Figure 2, black solid vertical line) using the peak of the F3 electrode since it presented the highest negativity compared to all other channels (see EEG topographies in Figure 2). Using a custom MATLAB (The MathWorks, Inc., Natick, MA, USA) tool (Küpper, 2012), we checked that every hand-marked position was shifted right to the peak of the maximum negativity at or close to F3 of each epileptic spike, whereby all markings were ensured to be at the same propagation phase of the epileptic activity. The continuous EEG and MEG data were then divided into spike epochs of 400 ms (200 ms before and after the spike peak). Finally, the EEG and MEG noise covariance matrices were calculated in the time range of -200 up to -50 ms before the spike peak.

**Figure 2.**
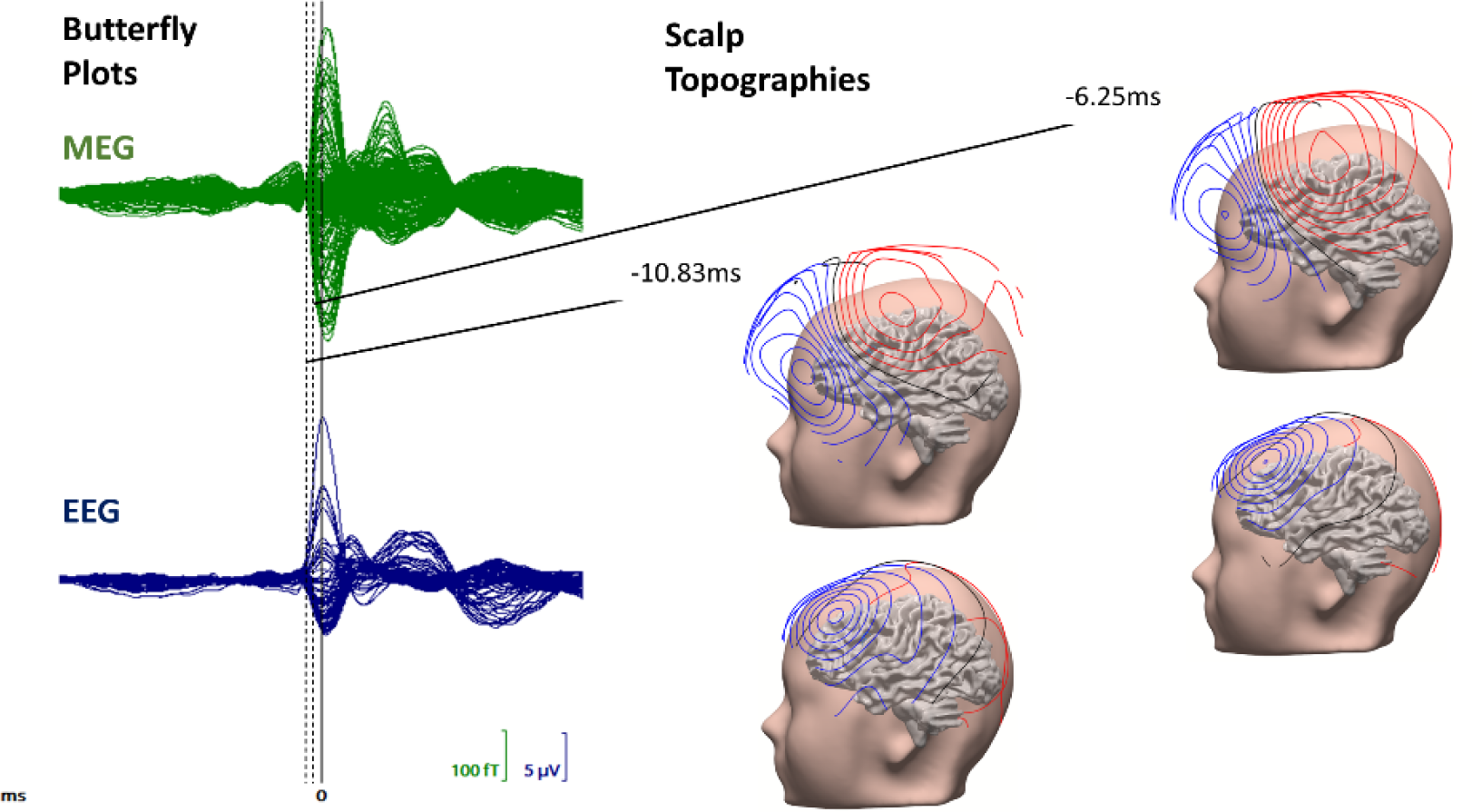
Grand-averaged MEG and EEG spikes: Butterfly plots (left) and topographies (right) of MEG (upper row, in green) and EEG (lower row, in blue) for the grand average across all 1050 individual spikes. The time points of interest are indicated with black-color vertical dashed lines. The scalp topographies are presented on the patient’s head model. The scalp and grey matter surfaces were generated in CURRY8 using the segmented images (see Section 2.4). The peak of the grand average is at 0 ms as indicated by the solid black vertical line in the butterfly plots. The presented amplitude range is in femto-Tesla (fT) and micro-Volt (μV) for MEG and EEG, respectively.

The butterfly plot of a grand average across all spikes is presented (Figure 2, left column) for MEG (in green) and EEG (in blue). The corresponding EEG and MEG scalp topographies (Figure 2, right columns) are presented for the selected time points, one at -6.25 ms, i.e., the middle of the rising flank of the grand average spike or the so-called *spike upstroke*, and at -10.83 ms, i.e., at the grand average *spike onset*.

### 2.4. Image Data Collection and Segmentation

A 3T scanner (MAGNETOM Prisma 3.0 T, Release D13, Siemens Medical Solutions, Erlangen, Germany) was used for the acquisition of MRI datasets. The measurement protocol was similar to (Aydin et al., 2017). We measured a 3D-T1-weighted (T1w) MRI using a fast gradient-echo pulse sequence with water selective excitation to avoid fat shift, a 3D-T2-weighted (T2w) turbo spin echo pulse sequence and a 3D-FLAIR turbo spin echo pulse sequence. We also measured diffusion tensor MRI (dMRI) using an echo planar imaging sequence, with one MRI scan with diffusion sensitivity b = 0 s/mm^2^ (i.e., flat diffusion gradient) and 20 MRI scans with b = 1000 s/mm^2^ in different directions, equally distributed on a sphere. Another MRI scan with a flat diffusion gradient, but with reversed spatial encoding gradients was acquired and used for susceptibility artifact correction (Ruthotto et al., 2012). During T1w-MRI measurement, gadolinium markers were placed at the same three landmarks as in MEG/EEG, namely, nasion, left and right preauricular points, for landmark-based registration of MRI to MEG/EEG.

CT images, acquired during the invasive video EEG evaluation to ensure correct electrode placement (see Section 2.2), contain a high-resolution representation of the cranial burr holes, which might affect the results of EEG source reconstruction (Benar & Gotman, 2002) and tDCS optimization (Datta et al., 2010; Sun et al., 2021). Therefore, these images were used to perform accurate modeling of these holes.

We used the T1w, T2w and CT images to construct an individual six-compartment (6C: scalp, skull compacta (SC), skull spongiosa (SS), CSF, gray matter (GM) and white matter (WM)) head model with four cranial burr holes (6CH). For this purpose, our segmentation approach includes additional image processing steps for an accurate segmentation of the cranial burr holes. We used the T1w-MRI for the segmentation of scalp, brain, and the three inner brain tissues (CSF, GW and WM). Subsequently, we first registered the T2w to the T1w and then produced a segmentation of the SC, CSF, and brain. The SS compartment was generated in two steps: (1) erosion of the full skull compartment to take into account a minimal thickness of the skull compacta and to define a region of interest (ROI) and (2) gray-value thresholding in this ROI in the T2w MRI. The SC compartment was then defined as the difference between full skull and the post-processed SS mask. All the segmented compartments were then post-processed to ensure that the masks did not overlap. Additional steps were necessary to model the burr holes in the skull using the CT images. Thus, we first registered the CT onto the T1w-MRI using FSL^1^. Then, we determined the SC and SS with holes, i.e., SCH and SSH, respectively, by segmenting the burr holes in the SC and SS compartments using the software Seg3D^2^. The location of the four holes was over the left frontal side of the skull bone. Each hole had a diameter of 6 mm on average. SCH and SSH were filled with material of scalp conductivity, assuming that one year after explantation of the invasive electrodes, the cranial holes are filled with collagenous tissue that has similar conductive properties as the scalp.

To model the white matter anisotropy in the realistic head model, the dMRI data was preprocessed, including an eddy current correction using FSL and a non-linear susceptibility artifact correction using both dMRI images with flat diffusion gradients (Ruthotto et al., 2012), as implemented in SPM12 as the HySCO subroutine^3^.

### 2.5. Generation of Realistic Finite Element Head Models

From the input six compartment (6C) segmented MRI, geometry-adapted hexahedral finite element method (FEM) meshes of 1 mm resolution were generated using SimBio-VGRID^4^ (Wagner et al., 2016). The corresponding FEM meshes have approximately 3.2 million elements and 3.3 million nodes. To model the WM anisotropic conductivity, we applied an effective medium approach (Rullmann et al., 2009) to calculate WM conductivity tensors from the preprocessed DTI data. The resulting FEM head volume conductor model then consists of six compartments with cranial burr holes and white matter tissue conductivity anisotropy (6CHA).

For the head model 6CHA_Cal, a calibration procedure for the individual estimation of skull conductivity was then carried out (Antonakakis et al., 2020). This was found to be of high importance because of a considerable inter-subject variability in skull conductivity (McCann et al., 2019; Antonakakis et al., 2020; Khan et al., 2023), which influences substantially EEG and EMEG source reconstructions (Dassios et al., 2007; Vorwerk et al., 2019). Finally, the conductivities for the compartments scalp, CSF and GM were based on standard literature-values (Antonakakis et al., 2020).

### 2.6. Source Analysis of Epileptic Activity using a Sub-Averaging Approach

We used the IED sub-averaging source reconstruction approach of (Aydin et al, 2015) that balances the number of averaged IEDs, and thus the resulting SNR, between i) large enough to avoid systematic error, especially in source depth, due to excessive residual data noise and ii) as small as possible to avoid losing the extent information of the underlying source area contained in the individual spikes, as would result from global averaging of all IEDs. This approach aims to identify the irritative zone in terms of localization, orientation, and extent of the underlying source patch from a set of sub-averaged spikes in EEG and MEG data. Each sub-average was reconstructed using a cortically-constrained deviation scan (CCDS) (Fuchs et al., 1998) on a source space with 2 mm resolution in the grey matter compartment of the individualized realistic head model. Within the inverse approach, FEM forward simulations for EEG and MEG were computed using the SimBio^5^ software. The CCDS results were further analyzed and visualized by means of its center of gravity, the so-called *centroid dipole*, and the surrounding *spread ellipsoid*. The approach was evaluated at the two time points of interest, the spike onset and the upstroke (Figure 2).

While we used all EEG electrodes for our sub-averaging and source reconstruction approach, for MEG, we gave more weight to the 123 gradiometers over the left frontal region-of-interest (ROI) to better focus on the bipolar MEG topographies spanned by those sensors (Figure 2), thereby increasing the goodness-of-fit of the CCDS, improving source reconstructions in the ROI and reducing the noise influence of especially the contralateral and occipital sensors far away from the ROI that mainly picked up noise.

To enable combined EEG and MEG source reconstruction, beside skull-conductivity calibration of the head model (Section 2.5), an SNR transformation was performed on both EEG and MEG converting them to unitless measurements (Fuchs et al., 1998). Each channel was whitened according to its individual noise level, which was estimated from the pre-trigger interval -200 ms to -50 ms.

As a final outcome, our sub-averaging procedure resulted in a centroid dipole and corresponding spread ellipsoid for each modality (EEG, MEG, EMEG). The EMEG centroid dipole was then used for targeted brain stimulation as presented in the following.

### 2.7. Multi-channel tDCS optimization

For an appropriate targeting of the EMEG centroid, a multi-channel tDCS (mc-tDCS) injected current should not only be maximal at the target area (intensity), and minimal at other areas (focality), but also predominantly oriented anti-parallel (inhibition) to the target orientation (directionality). Because of the complexity of the mc-tDCS targeting in the realistic head model, in the study at hand, we used for the first time our recently developed distributed constrained maximum intensity (D-CMI) optimization approach (Khan et al., 2022) in an epilepsy patient with the aim to non-invasively inhibit epileptic activity. D-CMI was evaluated in a somatosensory group study where it was shown to achieve nearly maximal target directionality, while reducing side effects and skin sensations and thus potentially improve sham conditioning (Kahn et al., 2023). Following the safety regulations (Antal et al., 2017), we used a total injection current of 4 mA and an injection limit per electrode of 2 mA. The D-CMI regularization parameter was chosen to make optimal use of the given 8 stimulation electrodes of our Starstim tDCS system (Starstim, Neuroelectrics, Barcelona, Spain). Due to the used Starstim neoprene cap we use 39 fixed positions of the tDCS electrodes on the scalp (standard 10/10 system) measured with the Polhemus device. The optimization scheme was applied two times: Initially, we performed D-CMI optimization using all 39 electrodes. Then, we selected those eight electrodes with the maximum absolute injected current and applied D-CMI again on only those.

Within the inverse optimization, SimBio-FEM was used for tDCS forward modeling (Wagner et al., 2014) in the 6CHA_Cal head model. The tDCS forward problem is closely linked to the EEG one, as Helmholtz reciprocity provides a common ground (Wagner et al., 2016; Gross et al., 2022). For a given mc-tDCS montage we first computed the electric potentials, from which the current density in all elements of the volume conductor was derived.

Finally, a quantification for the examination of the mc-tDCS optimization quality was done using three different measures (Khan et al., 2022), the *intensity in the target region* (IT in A/m^2^), the *intensity in the non-target regions* (INT in A/m^2^) and the directionality (DIR in A/m^2^), which measures the current intensity at the target side in the target orientation.

### 2.8. Study design of personalized and optimized mc-tDCS experiment

We first computed a D-CMI optimized mc-tDCS montage targeted to the EMEG centroid of our epilepsy patient (*D-CMI montage*). A second montage was created for sham-control, which uses the same 8 electrodes as the D-CMI montage, but minimizes brain stimulation, while preserving the burning and tingling sensation at the skin level, similar to the real stimulation (*Active-sham* montage). We used a double-blind experimental setup, i.e., both the patient as well as the treating physician, medical technical assistants and nurses were fully blinded regarding the two stimulation conditions. The management and control software for mc-tDCS was set up by independent research staff from the Institute for Biomagnetism and Biosignalanalysis beforehand and stimulation parameters were hidden to all other participants in the experiment. The main goal of this double-blind sham-controlled N-of-1 trial was thus to investigate the effect of our non-invasive personalized D-CMI stimulation on IED frequency. Our hypothesis is that D-CMI inhibits the irritative zone and thus reduces the frequency of IEDs in EEG’s measured after stimulation when compared to those measured before, while sham has no effect on IED frequency.

An overview of our experimental setup is shown in Table 1. We stimulated the patient twice per day for 20 minutes each, with a pause of 20 min in between, because 2 × 20-min daily stimulation was found to be superior to the protocol using 20-min daily stimulation only (Yang et al., 2020). Due to the circadian rhythm of IEDs (Kaufmann et al., 2021; Baud et al., 2018), we always stimulated between 11 and 12 AM. Each stimulation condition was applied for 5 days in a single week, with a washout period of 5 weeks between the two stimulation weeks to avoid any interference or carry over effect (Woods et al., 2016). One-hour blocks of EEG were measured directly before and after the stimulation block every day. Before the first stimulation in each stimulation week, 2 hours of EEG were measured for better baseline IED detection, while in the analysis, the marked IEDs in the two-hour block were directly averaged, unless otherwise stated. Those measurements were acquired by a clinical EEG system (Nihon Kohden, Tokyo, Japan) with 19 electrodes (subset of 10-10 system) and a sampling frequency of 200 Hz. Additionally, our sham condition was also used to control for unknown underlying ultradian rhythms that may be out of sync between days (Spencer et al., 2016).

**Table 1.**
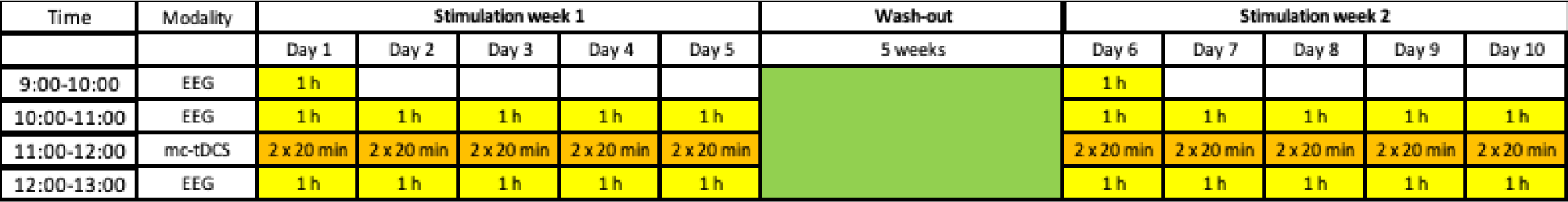
tDCS experimental setup: In two stimulation weeks, separated by a wash-out phase of 5 weeks (in green), either real stimulation using the D-CMI montage or sham-stimulation using the Active-sham montage are applied twice every weekday for 20 min each, with a 20 min pause in between (in orange). Before and after stimulation (in yellow), 1 h of EEG is measured, which is later marked for IEDs by three independent epileptologists. On the first day of a stimulation week, 2 h of EEG are measured to better determine the IED baseline level.

Standard questionnaires from the latest reviews for tDCS (Antal et al., 2017) and specialized questionnaires adapted to the patient’s specific form of focal epilepsy to assess tolerability and symptoms during and after mc-tDCS using a 5-point rating scale (1 = no sensation, 5 = extreme sensation) as well as relevant patient information were used.

### 2.9. IED detection in mc-tDCS EEG data and statistical evaluations

The IED marking in the pre- and post-mc-tDCS stimulation EEG data was performed by three experienced epileptologists (co-authors SR, CK, SC). The EEG data files were cut into 1-hour segments, completely anonymized, and labeled with code names (FK), so that the epileptologists had no information about time or order, to keep them completely blinded during the manual annotation.

We used a mixed ANOVA to evaluate influence of Pre/Post, type of treatment and expert marker (Wilson et al., 2016) on spike frequency and between days using SPSS^6^. Assumptions of normality were tested with Shapiro-Wilk while homoscedasticity was investigated with Levene’s Test. Sphericity on the other hand was given, since the within-days factor Pre/Post only had two steps. Post-hoc multiple comparisons were elucidated by Tukey’s HSD (honestly significant difference) test. Then, we considered that the measurements of IED frequency before and after any given stimulation (D-CMI or Active-sham) block should be paired (or dependent) while data measured on different days should be considered independent. Our between-days factor was then considered the type of treatment” (D-CMI or Active-sham).

### 2.10. Invasive EEG diagnosis and neurosurgical procedure

A second invasive EEG diagnosis with depth electrodes (iEEG) was performed after the tDCS experiment in autumn 2021. The implantation of depth brain electrodes was again performed in general anesthesia using a stereotactic frame (Leksell Frame G, Elekta). Craniotomy and resection under awake condition then followed in January 2023.

## 3. Results

We divide our results section into two subsections. In the first subsection 3.1 we investigate source analysis differences between EEG, MEG and combined EEG/MEG, co-localize these source reconstruction results with 3D-FLAIR MRI and with a second iEEG investigation and thereby increase confidence in the reconstruction of the patients’ epileptogenic zone close to Broca’s area. Subsection 3.2 then presents the results of our non-invasive transcranial electric stimulation experiment of Table 1, where we show that individually targeted and optimized mc-tDCS significantly inhibited epileptiform activity, while Active-sham did not. Figure 3 schematically depicts our proposed approach from the measured non-invasive neuroimaging data over the target source reconstruction and diagnosis to the targeted and D-CMI optimized montage calculation.

**Figure 3.**
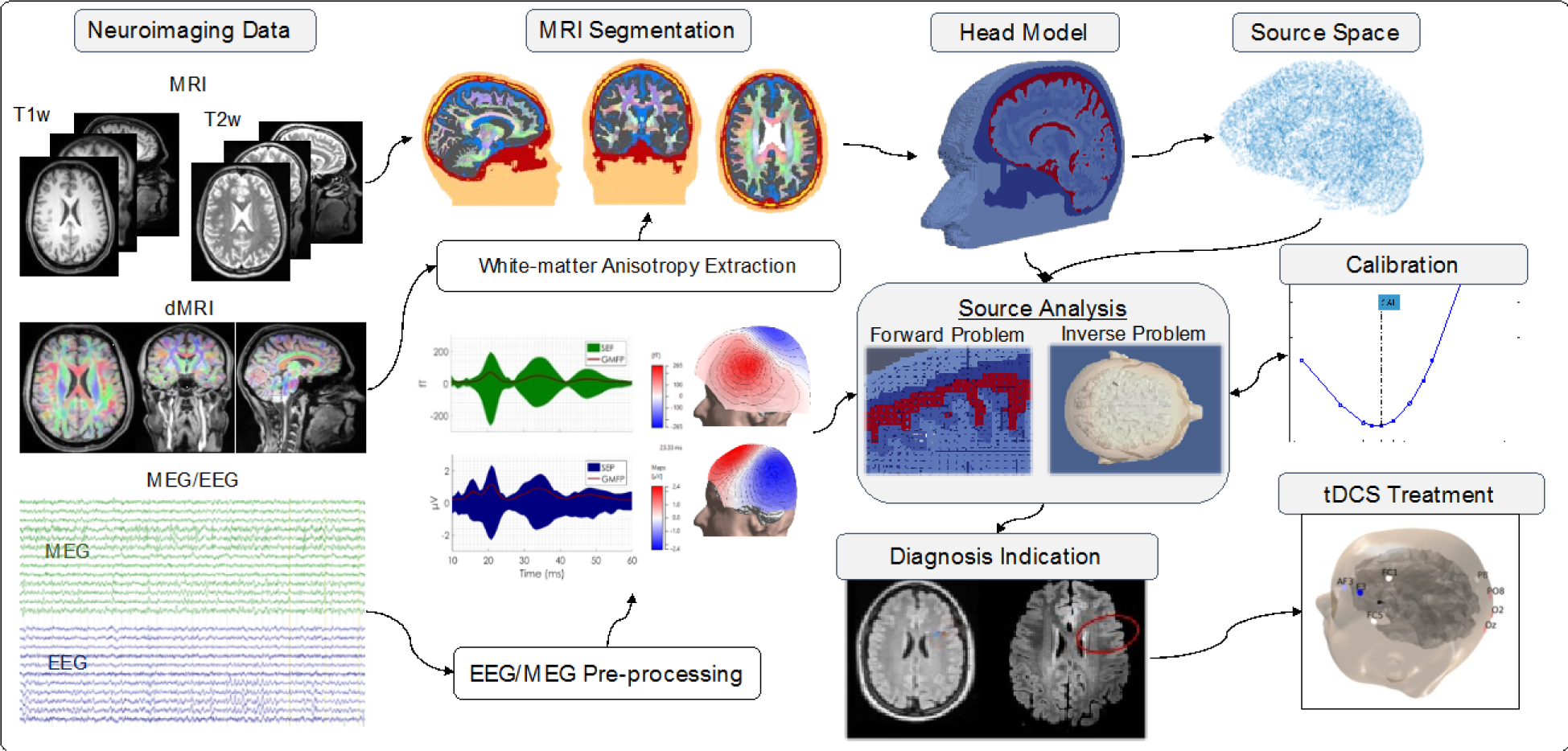
Flowchart for epilepsy diagnosis and tDCS treatment: A flowchart from the neuroimaging data (MRI, diffusion MRI (dMRI), and EEG/MEG) over the EEG/MEG preprocessing, MRI segmentation and diagnosis by source analysis and FLAIR-MRI to the D-CMI optimized mc-tDCS treatment is presented.

### 3.1. Combined EEG/MEG Source Reconstruction for the Identification of a cortical malformation

#### 3.1.1. Results of Sub-Averaging Source Reconstructions

Our calibration procedure (Section 2.5) resulted in skull conductivities of 0.0041 S/m (0.0148 S/m) for compartments SSH (SCH), respectively, which were then used in the calibrated head volume conductor model, 6CHA_Cal.

Following the sub-averaging strategy of Section 2.6, we performed source reconstructions at the two time points of interest, i.e., the spike onset (-10.83 ms) and the spike upstroke (-6.25 ms) for the combined EEG/MEG, single modality EEG and single modality MEG. The corresponding source reconstructions are presented in Figure 4 (centroids with spread ellipsoids) and Table 2 describes the source reconstruction differences between the different modalities with regard to centroid location, orientation and strength. Regarding location, centroids and spread ellipsoids are in or near Broca’s area, i.e., Brodmann areas 44 (pars opercularis) and 45 (pars triangularis). EMEG locations are more influenced by MEG than by EEG. Regarding source orientations, EEG dipoles show rather radial and MEG dipoles rather tangential orientations, while EMEG orientation is influenced more by EEG than by MEG.

**Figure 4.**
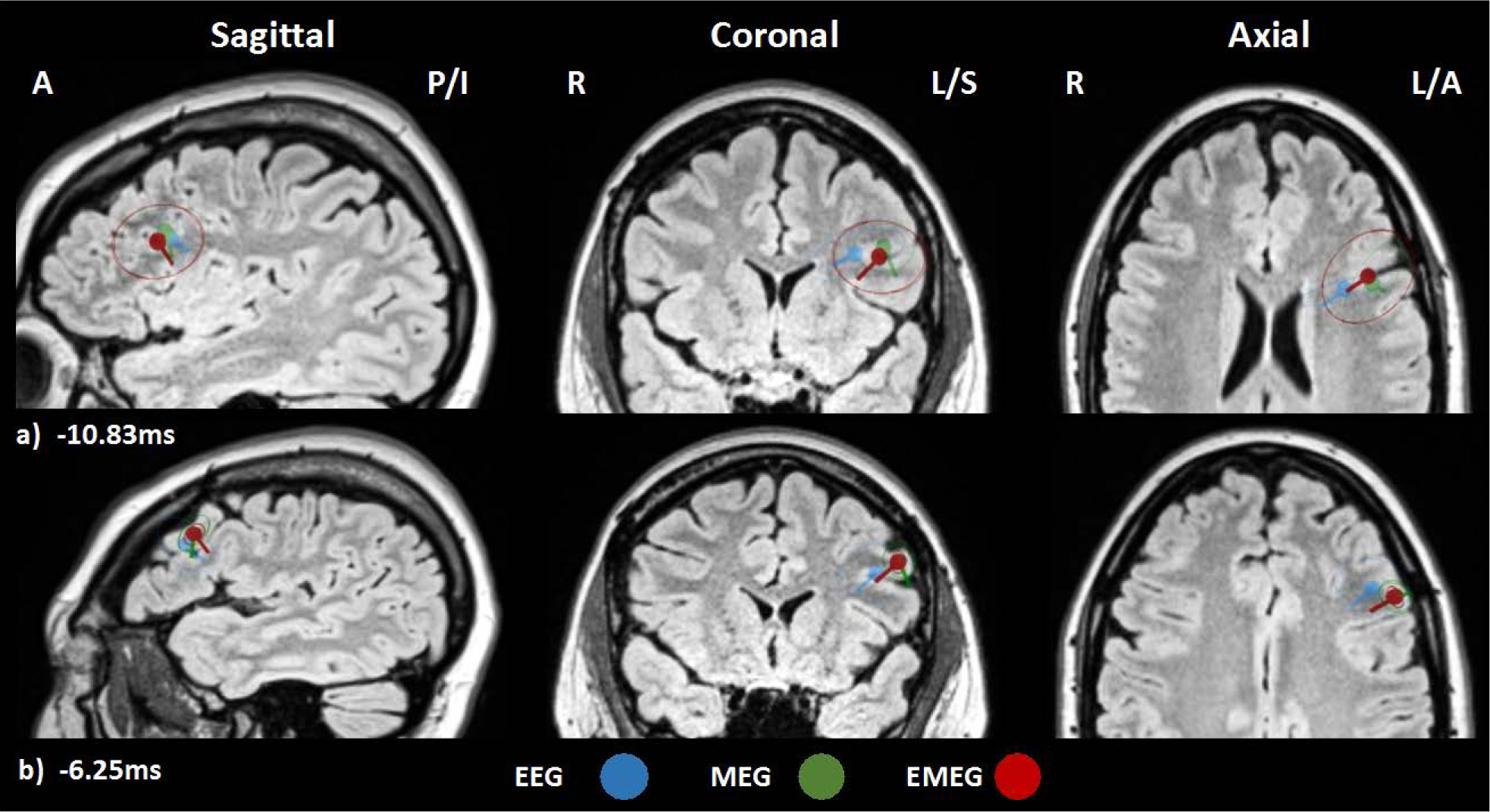
Centroids and spread ellipsoids for the two time-points and three measurement modalities: Spike cluster centroids and spread ellipsoids are depicted on 3D-FLAIR MRI cutplanes (sagittal (left column), coronal (middle column) and axial (right column)) at the two time-points **a)** -10.83 ms (spike onset) and **b)** -6.25 ms (spike upstroke) for the measurement modalities EEG (blue), MEG (green) and EMEG (red). Cutplanes are centered at the position of the EMEG centroid. The size of all centroids is kept constant. A: anterior, P: posterior, I: inferior, R: right, L: left and S: superior.

**Table 2.** Differences in centroid reconstruction at spike onset and upstroke with regard to measurement modality (EEG, MEG, EMEG) for head model, 6CAH_Cal: Differences in centroid location (third column), orientation (fourth column) and strength (fifth column) between measurement modalities (second column) at the latencies of interest (first column).

| Latency (ms) | Modality Pair | Location Diff. (mm) | Orientation Diff. (degree) | Strength Diff. (%) |
| --- | --- | --- | --- | --- |
| -10.83 (-6.25) | EMEG vs. EEG | 9.6 (9.3) | 18.7 (8.1) | -14.3 (-13.9) |
|  | EMEG vs. MEG | 4.2 (0.6) | 66.9 (73.8) | 57.1 (64.2) |
|  | EEG vs. MEG | 10.3 (9.7) | 82.5 (74.1) | 62.4 (68.6) |

At the spike onset (Figure 4a), all centroids are localized in deep areas of the left frontal cortex. The EMEG centroid (Figure 4a, in red) is located at the bottom of the sulcus between EEG and MEG centroid with distances of 9.6 mm and 4.2 mm, respectively. The orientation of the EMEG centroid is more influenced by EEG (remaining angular difference: 18.7°) than by MEG (remaining angular difference: 66.9 °), as the radial orientation component pointing into the sulcus is stronger, even though a smaller tangential orientation component from the right sulcal wall has been contributed by the MEG. With regard to the centroid strengths, EMEG is slightly smaller than EEG (- 14.3%), while it is considerably larger than MEG (73.8%).

At the spike upstroke (Figure 4b), all centroid dipoles are localized to more superficial positions at the gyral crown in the left frontal cortical region. There is thus a considerable amount of propagation from spike onset to upstroke. With a distance of 0.6 mm, the EMEG centroid is at nearly the same location as the MEG one, while with 9.3 mm the distance to the EEG one is larger. The reverse is true for centroid orientation and strength, because the EMEG centroid orientation and strength is influenced more by the EEG (only 8.1 degrees orientation and 13.9% strength difference between EMEG and EEG centroids, see Table 2) than by the MEG (73.8 degrees orientation and 64.2% strength difference between EMEG and MEG centroids, see Table 2).

When comparing EEG and MEG, we find considerable differences (Table 2, third row). At the spike onset (spike upstroke), the EEG and MEG centroid locations differ by 10.3 mm (9.7 mm), while the orientation components are even nearly orthogonal to each other (82.5° and 74.1 ° orientation differences for spike onset and upstroke, respectively, see Table 2). As the EEG centroids show higher strengths than the MEG centroids at the spike onset (62.4%, see Table 2), there is evidence that the mainly radially oriented pyramidal cells at the bottom-of-sulcus build the larger part of the overall epileptic patch when compared to the mainly tangentially oriented pyramidal cells at the neighboring right side of the sulcus wall (see right upper subfigure in Figure 4a). With 68.6%, the situation is similar at the spike upstroke. This interpretation is also supported by the rather radial source topographies of EEG and rather tangential source topographies of MEG at spike onset and upstroke in Figure 2.

The quality of the source reconstruction depends on the SNR and the GOF, which in turn will also influence the spread ellipsoids. As Figure 4 shows, due to the lower signal-to-noise ratio (SNR) at the spike onset (EEG: 2.67, MEG: 2.61 and EMEG: 2.6; see Figure 2), the spread ellipsoids are larger than with the higher SNR at spike upstroke (EEG: 4.5, MEG: 6.1 and EMEG: 5.6; see Figure 2).

A similar trend can be observed for the GOF, which is lower at the spike onset (EEG: 80%, MEG: 74% and EMEG 82%) when compared to the spike upstroke (EEG: 91%, MEG: 85% and EMEG: 82%). Due to the need for regularization of MEG in a realistic head model (spatially high-frequent data noise might otherwise be amplified in large and unrealistic radial orientation components), lower GOF values are achieved for MEG and EMEG when compared to EEG, where no regularization is used (Fuchs et al., 1998).

In summary, in this section we reconstructed the EMEG centroid and spread ellipsoid at spike onset (Figure 4a, in red), which will serve in the following to guide (i) a retrospective re-evaluation of 3D-FLAIR MRI and (ii) a second invasive EEG with the overall goal to increase the confidence in the determination of the epileptogenic zone.

#### 3.1.2. Detection of cortical malformation and seizure onset area

Combined EEG/MEG might allow reconstruction of also deeper sources at the spike onset (Aydin et al., 2015,2017), where the epileptic activity might not yet be subject to propagation, in contrast to spike upstroke (Aydin et al., 2015,2017) and especially spike peak (Lantz et al., 2003). We thus supposed that the EMEG centroid and corresponding spread ellipsoid at spike onset (Figure 4a), localized at the bottom of a sulcus in the left frontal cortex next to Broca’s area and thus matching the patient’s seizure semiology may represent the origin of the epileptic activity. We then co-registered the EMEG centroid at spike onset to the patient’s 3D-FLAIR MRI (Figure 5). At the region-of-interest (ROI), the cortex was thickened and the grey-white matter boundary was slightly blurred, suggesting a cortical malformation, which had not been detected beforehand (Figure 5b, area in red circle).

**Figure 5.**
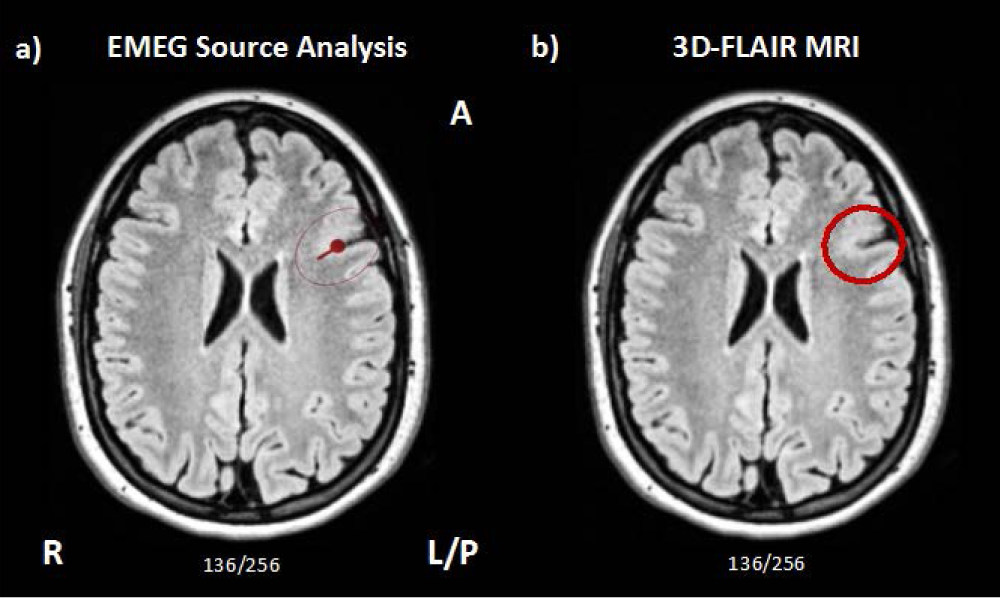
3D-Flair MRI: Evidence of cortical malformation in region-of-interest defined by EMEG source analysis: **a)** EMEG source reconstruction at the spike onset (-10.83 ms) (red centroid with corresponding spread ellipsoid). **b)** Deep sulcus with thickened cortical band and blurred grey-white matter junction, suggesting a cortical malformation, which had not been detected beforehand (center of the red circle in the axial cutplane through the EMEG centroid location of the patient’s 3D-FLAIR MRI). The slice number is indicated at the bottom of each image. A: anterior, P: posterior, I: inferior, R: right, L: left and S: superior.

Due to the converging evidence of seizure semiology, EMEG source reconstruction at spike onset and the retrospectively suspected cortical malformation in the 3D-FLAIR MRI, a second video-EEG with invasive EEG electrodes was performed in 2021 (Figure 6), with the implantation guided by the EMEG centroid result at spike onset. This second iEEG study then confirmed that the seizure onset zone is located in the area of the suspected cortical malformation.

**Figure 6.**
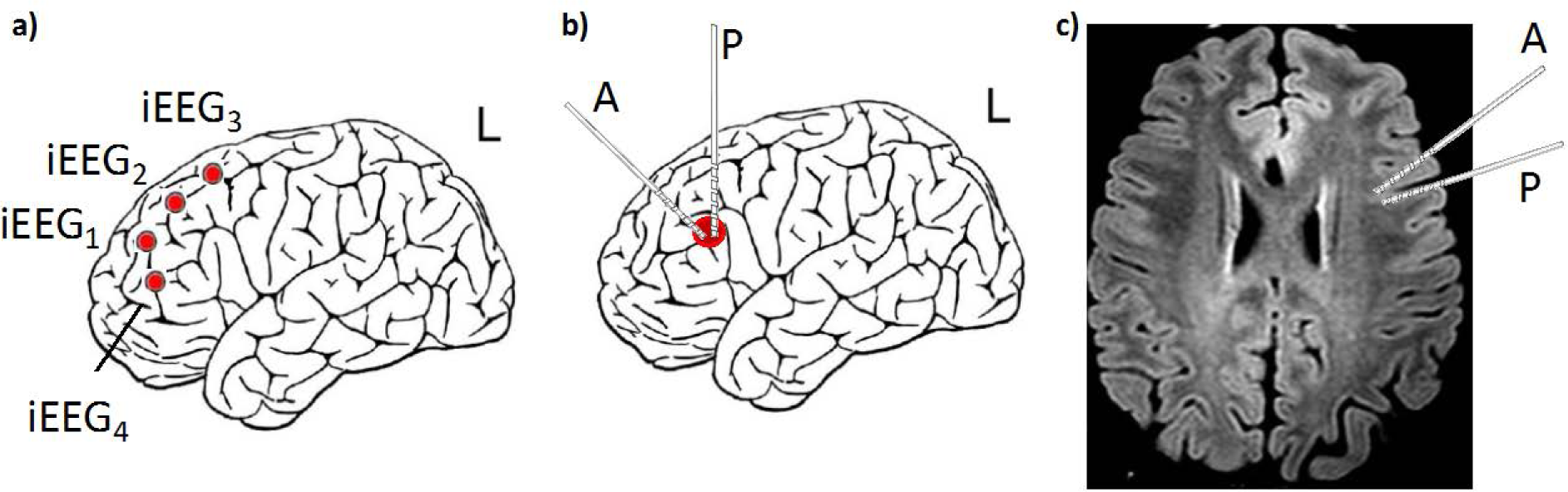
Comparison of the two invasive EEG evaluations: **(a)** First iEEG from 2018, see also Figure 1; (b,c) Second iEEG from 2021 with depth electrode implantation guided by the EMEG source analysis and 3D-FLAIR MRI results. The seizure onset zone could be located to the suspected area of cortical malformation at the spike-onset EMEG centroid from Figure 5a. A: anterior, P: posterior, L: left.

### 3.2 EMEG-targeted and D-CMI optimized mc-tDCS montage

For the EMEG centroid at the spike onset as target (Figure 5a; black cone in Figure 7b), we used D-CMI optimization to compute a personalized 8-channel mc-tDCS montage (Figure 7b). In Figure 7a, we present the resulting electric field and current distribution over the cortical surface and inside the target area, showing the high directionality for inhibiting the seizure onset zone as well as the reasonable focality of our D-CMI approach, distributing the anodal currents over occipital areas of the head (Figure 7b).

**Figure 7.**
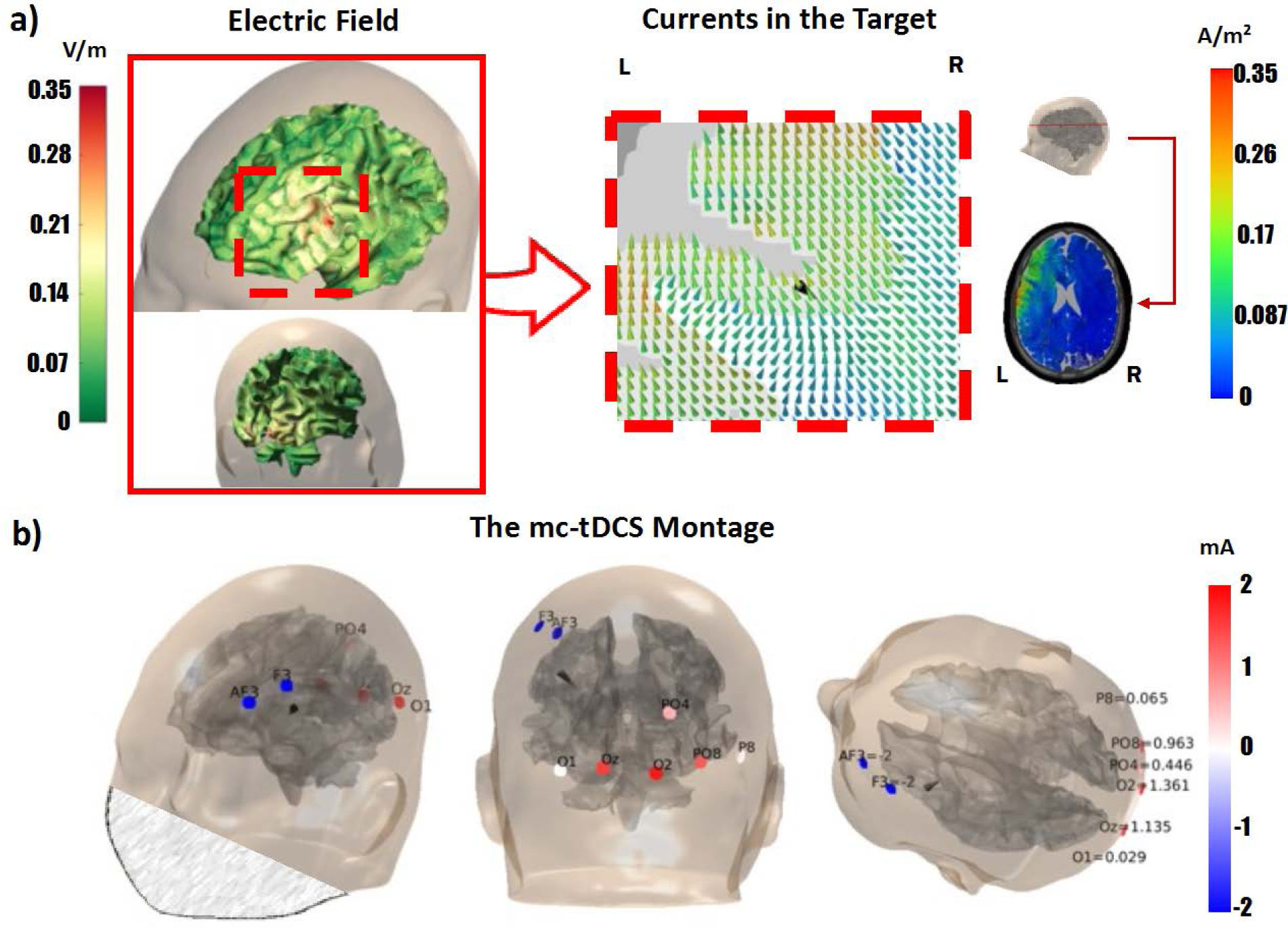
EMEG targeted and DCMI optimized 8-channel mc-TDCS montage. **a)** Distribution of the simulated electric field over the patient’s cortex (left subfigure) and zoomed view of the injected current (color-coded cones) in the target region (black cone) (right subfigure). **b)** Three different views show the D-CMI optimized 8-channel mc-tDCS montage, targeted at the EMEG centroid at spike onset, used for the D-CMI stimulation condition. The sum of absolute values of all currents is 8 mA with a limitation of max 2 mA per electrode. The patient’s face has been blurred in a) and b) for anonymization purposes.

Secondly, for sham-control, we created an Active-sham stimulation montage using the same most important four electrodes as in D-CMI, but changed currents, namely AF3 and Oz as anodes with -2 mA and F3 and O2 as cathodes with +2 mA. As a result, the injected currents at the anodes AF3 (Oz) are directly extracted at the neighboring cathodes F3 (O2), leading to a considerably lower simulated DIR (D-CMI: 0.21 A/m^2^ / Active-sham: 0.03 A/m^2^), IT (D-CMI: 0.3 A/m^2^ / Active-sham: 0.04 A/m^2^) and INT (D-CMI: 0.09 A/m^2^ / Active-sham: 0.01 A/m^2^). The Active-sham montage thus minimizes brain stimulation while preserving the burning and tingling sensation at the skin level in the left frontal and right occipital area, just like the real stimulation does. We tested this before patient stimulation with the experimenters as subjects, who were not able to distinguish the two stimulation conditions.

### 3.3 Double-blind sham-controlled mc-tDCS experiment

Figure 8 visualizes the amount of interictal epileptiform discharges (IEDs) before (Pre) and after (Post) either optimized tDCS (left) or Active-sham stimulation (right) by our three epileptologists (different green colors, average of all three in blue).

**Figure 8.**
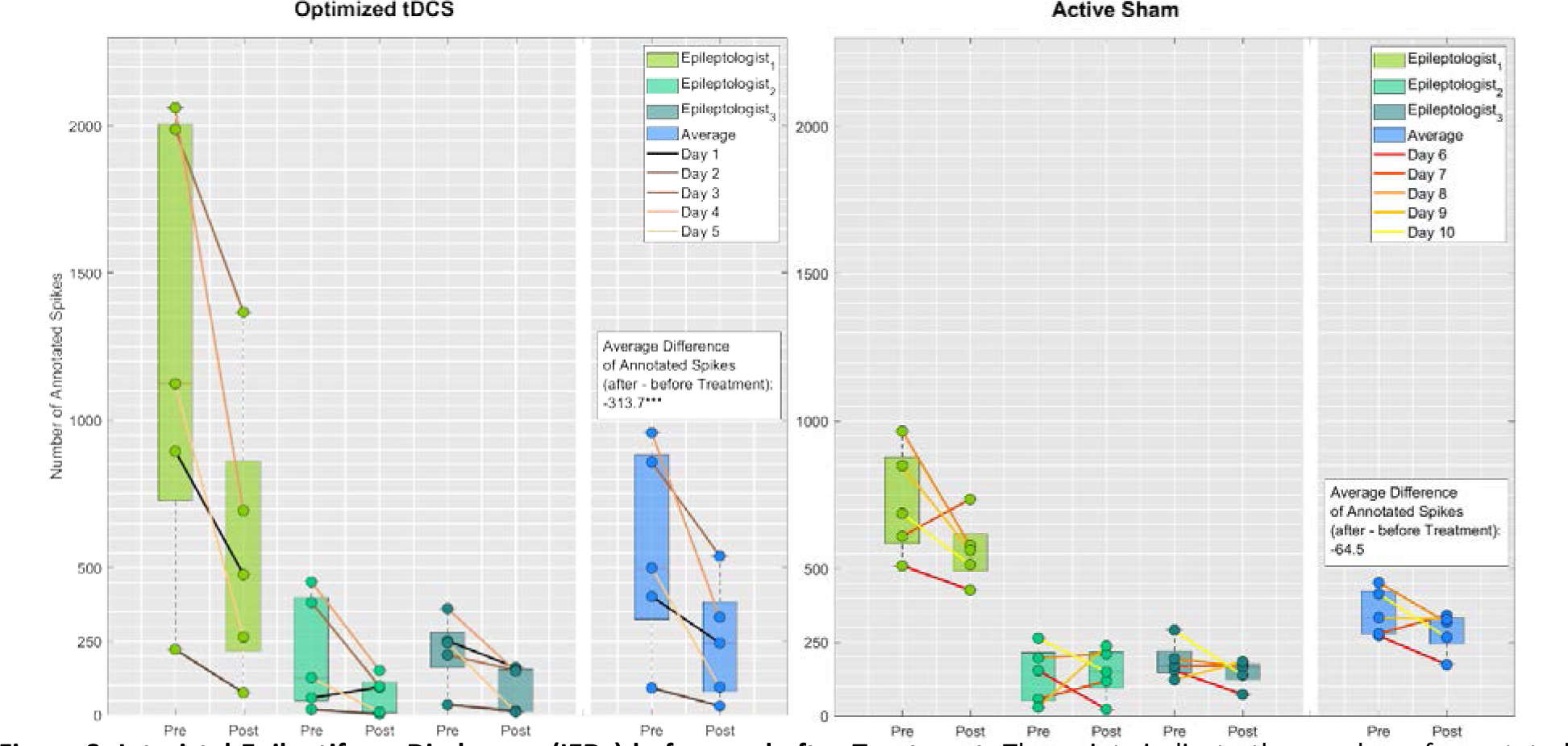
Interictal Epileptiform Discharges (IEDs) before and after Treatment. The points indicate the number of annotated spikes (marked IEDs) during 1 hour of EEG found by Epileptologist 1 (light green), 2 (medium green), 3 (dark green) and their average (blue) before (Pre) and after (Post) treatment. Boxplots show the median (central mark) and the 25th and 75th percentiles (box), and the whiskers extend to the most extreme data-points which are not considered to be outliers. **Left:** Treatment is D-CMI optimized mc-tDCS targeted by the EMEG centroid at spike onset. The average change in Incidence of IEDs through treatment is -313.7 IEDs per hour (p<0.0001). **Right:** Treatment is Active Sham. The average change in Incidence of IEDs through treatment is -64.5 IEDs per hour (p>0.05).

We find a statistically significant main effect of Pre/Post on IED frequency (F(1.26)=18.18, p<0.001), a statistically significant interaction between Pre/Post and expert marker (F(2.26)=6.9, p<0.01) and a statistically significant interaction between Pre/Post and type of treatment (F(1.26)=7.89, p<0.01).

The only group of data that saw its assumption of normality being rejected was due to an outlier at day 1 (black curve in Figure 8) for Epileptologist_2_ during the first stimulation week. Here, in contrast to all other markings, the IED frequency is elevated after stimulation. Mixed ANOVA is quite robust in terms of a rejected normality assumption, especially for only a small part of the overall data. It was therefore decided to keep the outlier in. Levene’s test confirmed homoscedasticity between groups for every step of our within-days factor Pre/Post.

Because the interaction type of treatment with Pre/Post is disordinal, the main effect of Pre/Post on IED frequency cannot be sensibly interpreted (Pedhazur & Schmelkin, 1991).

Averaged across all epileptologists (blue in Figure 8), according to the Tukey-HSD, D-CMI leads to a highly significant reduction in IED frequency with -313.7 IEDs per hour (p < 0.0001), while Active-sham did not (p > 0.05) (Figure 8). Reductions over the 5 days of real stimulation are between 37% and 81% (mean±SD: 58% ±19%), while they are between -23% and +36% (mean±SD: 16% ± 26%) for Active-sham. Side effects were exclusively transient sensations. The only detected seizure occurred during the stimulation week but had decreased severity with preserved awareness and recall of the conversation going on immediately before the seizure.

Questionnaires showed that the patient could not distinguish D-CMI and Active-sham. No adverse effects occurred during or after D-CMI or Active-sham and none of the treatment blocks had to be cancelled. According to the questionnaire which used a 5-point rating scale (1 = no sensation, 5 = extreme sensation), the patient reported itching on the back of the head during D-CMI (Active-sham) stimulation with a mean intensity of 3.4±0.55 (2.8±0.45). Further sensations were pain under electrode F3 with 4.2 ± 0.84 (3.4 ± 0.55), burning with 2 ± 1 (2 ± 1.41), warmth 2.2 ± 0.84 (1.4 ± 0.89), tiredness or decreased attention 1 ± 0 (1 ± 0) and dizziness 3.2 ± 0.45 (1 ± 0). Only dizziness differed significantly in intensity between D-CMI and Active-sham (p<0.001).

### 3.4 Outcome of epilepsy surgery

Due to the location of the seizure onset zone close to the motor language area (Broca’s area) and arcuate fasciculus, we decided to perform craniotomy and resection under awake condition. After the neurosurgical intervention in January 2023, the patient did not show any neurological deficits and is seizure-free under medication. Furthermore, the postoperative MRI confirmed the correct location of resection.

## 4. Discussion

In this N-of-1 trial, we applied for the first time individually targeted distributed constrained maximum intensity (D-CMI) optimized multi-channel transcranial direct current stimulation (mc-tDCS) in a patient with refractory focal epilepsy. The goal of reducing epileptic activity non-invasively was successfully achieved, as shown by a double-blinded sham-controlled stimulation experiment. The success of the optimization is dependent on the targeting accuracy with regard to both target location and target orientation (Dmochowski et al., 2013; Zulkifly et al., 2022; Khan et al., 2023) as well as on the idiosyncrasies of the individual head volume conductor model (Datta et al., 2010; Kasten et al., 2019; Khan et al., 2023; Beumer et al., 2022). Here, we targeted by combined EEG/MEG (EMEG) source analysis using the different sensitivity profiles and complementary information contained in both modalities (Dassios et al., 2007; Aydin et al., 2017; Ebersole and Wagner, 2018; Duez et al., 2019; Piastra et al., 2021). For EEG, MEG and mc-tDCS electromagnetic field modeling, we used the finite element method (FEM) in a realistic anisotropic six-compartment head model calibrated for individual skull-conductivity (Antonakakis et al., 2020; Khan et al., 2023) and taking into account cranial burr holes from a previous invasive EEG (Benar & Gotman, 2002; Lau et al., 2016; Datta et al., 2010). To the best of our knowledge, this level of personalization in targeting, optimization and head modeling for non-invasive diagnosis and therapy of focal epilepsy has not yet been proposed and used before. The most important finding of our mc-tDCS experiment is that, when comparing the IEDs marked by the three epileptologists in the EEGs before and after stimulation, we find a statistically highly significant (p<0.0001) reduction of IEDs in the D-CMI stimulation week, while this is not the case for Active-sham (p>0.05) (Figure 8). On average across the three epileptologists, the percentage reduction of IEDs between Pre and Post stimulation for the five days of stimulation per condition are always higher for D-CMI (between +37% and +81%) (mean ± SD: 58 ± 19%) than for Sham (between -23% and +36%) (mean ± SD: 16 ± 26%), where in the latter it also varies between reduction and increase (Figure 8, in blue). When compared to (Kaufmann et al., 2021), who reported reductions of IEDs of mean 30% with SD of ± 21% (p = 0.001), we thus achieved a high effect size with our individualized therapy.

(Baud et al., 2018; Kaufmann et al., 2021) showed circadian and multidian rhythms for IED frequencies with different chronotypes (Figure 3b, c, d in Baud et al., 2018) and differences between neocortical and mesiotemporal epilepsies (Figure 3a in Baud et al., 2018). A purely circadian rhythm cannot explain our data because the read-out EEG blocks were separated by only the 1 hour of stimulation, always from 11:00 to 12:00AM (Table 1), and the presented highly significant reduction of IEDs shown in the real stimulation week is accompanied by a concomitant lack of reduction in IEDs in the sham stimulation week. On the other hand, the overall lower level of IEDs in the second (sham) stimulation week cannot be explained solely by the long-term stimulation effect of our real stimulation, performed six weeks earlier. At least in part it may also be explained by the multidian rhythm of 26 days from peak to peak, corresponding to 39 days from peak to trough of IED frequency (Figure 1f in Baud et al., 2018), as there were 42 days (6 weeks) between our two stimulation weeks (Table 1). Following the results of (Baldin et al., 2017; van Campen et al., 2016), another explanation could be the patient’s stress level, which was higher in the first (shortly before final training examinations) than in the second stimulation week (after exams). In addition to these factors, IED frequencies may have been modulated by sleep-wake cycle (Vignatelli et al., 2010) and environmental factors (Rakers et al., 2017), which may have contributed to the strong daily IED fluctuations seen in our data (Figure 8).

(Karoly et al., 2016) showed that the relationship between IEDs and seizures is subject-specific and that it is not clear that spikes directly promote or inhibit seizures. However, it was hypothesized that spikes and seizures share one or more common regulatory mechanisms, as evidenced by their similar circadian patterns. In the case of our patient, who suffered only a single seizure of reduced severity on the evening of the second day of the real stimulation week, it is interesting to note that the average IED frequency was reduced on the first stimulation day from 402 down to 245 and on the second day from 92 down to 31, before the seizure occurred, increasing the pre-stimulation IED baseline to 858 on the third day.

As shown in this study and supported by the group studies in healthy subjects of (Khan et al., 2023,2022), D-CMI optimizes for (i) high directionality (DIR), i.e., current amplitude anti-parallel (for inhibition) to the target vector orientation and thereby also to (ii) high target intensity (IT). It also reduces (iii) skin sensations and (iv) non-target intensity (INT) and thereby uncontrolled side-effects. For our inhibition goal, DIR is a particularly important measure. It is part of the D-CMI functional to be maximized (formula (2) in Khan et al., 2022) and includes aspects of both target location and orientation, while most other studies focus on target location only (cathode over target, anode far away) (Ashrafzadeh et al., 2023; Kaufmann et al., 2021; Yang et al., 2020; Boon et al., 2018). However, for focal targets, the inclusion of target orientation has been shown to contribute considerably to higher effect sizes and control over stimulation effects by reducing the often reported high inter-subject effect variability (Kahn et al., 2023; Evans et al., 2022; Zulkifly et al., 2022; Seo et al., 2017; Krieg et al., 2015), while for non-focal targets, optimizing for highest IT might be in the foreground (Dmochowski et al., 2013). Our mc-tDCS experiment was sham-controlled, i.e., the real D-CMI stimulation condition was accompanied by an Active-sham condition in order to improve blinding (Neri et al., 2020). For our Active-sham, we used the same stimulation electrodes as in D-CMI, but a different current injection pattern, optimized for the same sensations at the skin level, but low IT, INT and DIR measures. The patient questionnaires showed that the same sensations at the skin level were achieved, as the patient was not able to distinguish Active-sham from real D-CMI stimulation. In our FEM simulation using the realistic head volume conductor model the Active-sham stimulation leads to a 6.7 times lower DIR (and a 7.4 times lower IT) than D-CMI. In combination with our experimental results of Figure 8, we can therefore assume that our sham stimulation was effective. We furthermore used a double-blind experimental design in which both (i) the patient and (ii) the treating personnel, i.e., the three epileptologists who marked the IEDs, the medical technical assistants, and the nurses, were fully blinded regarding the two stimulation conditions.

Our somatosensory study in a group of healthy subjects indicated that EMEG targeted and D-CMI optimized mc-tDCS can outperform standard bipolar stimulation (Khan et al., 2023). In this clinical investigation, we therefore decided against a third bipolar stimulation condition because of the already challenging experimental design, expecting higher effect size, less side-effects and skin sensations and more control over the stimulation outcome of our highly individualized mc-tDCS procedure (Khan et al., 2023). While standard bipolar montages (cathode over target area) have also been successfully used to reduce epileptic activity in focal epilepsy (Ashrafzadeh et al., 2023; Kaufmann et al., 2021; Yang et al., 2020), their targeting is ignoring target orientation and, additionally, is also often suboptimal with regard to target location, overall achieving a sub-optimal DIR (Khan et al., 2023). However, bipolar montages could also be individually optimized for DIR. In our patient, following Helmholtz’ reciprocity (Gross et al., 2021; Khan et al., 2023), we could have positioned the cathode at the potential trough of the EEG spike onset topography (Figure 2) between F3 and AF3 (Figure 7) and the anode at the potential peak at electrode O2 (Figures 2 and 7). Such a bipolar montage would have even been the result of an EMEG targeted maximum intensity (MI) optimized mc-tDCS (Khan et al., 2023), it would thus have optimal DIR. However, patients would hardly tolerate current amplitude of 4 mA through just a single electrode without local anesthesia and, more importantly, D-CMI achieves a better balance by distributing over multiple electrodes, especially on the side far from the target (in our case, the occipital anodes), thereby increasing focality and reducing side-effects as well as skin sensations, while almost maintaining the high directionality of MI (Khan et al., 2023).

The reduction of skin sensations by D-CMI enabled an overall 4 mA of injection current (due to the distribution over the electrodes with a maximum of 2 mA per electrode), while recent studies used only 2 mA (Kaufmann et al., 2021; Yang et al., 2020). One reason for our higher mean in reduction of IEDs (58% versus 30% of (Kaufmann et al., 2021)) may thus also be the higher current magnitude we were able to use due to D-CMI optimization.

A second main result of our work is that the EMEG centroid at spike onset (Figures 4a,5a) was localized at the bottom of a sulcal valley in the left frontal lobe close to Broca’s area, in concordance with the patient’s seizure semiology. This EMEG localization guided a retrospective re-evaluation of FLAIR-MRI (Figure 5b) and a subsequent iEEG (Figure 6b,c), leading overall to a successful localization of the epileptogenic zone, thereby supporting the neurosurgery. Single modality EEG or MEG were sensitive to different aspects of the extended source patch, EEG to the radially-oriented sources at the sulcal valley and MEG to the tangentially-oriented ones at the right sulcal wall. At spike upstroke, sources may have already propagated from the bottom-of-sulcus area close to the cortical malformation (Figure 4a) to more superficial gyral crown areas (Figure 4b). This propagation phenomenon is well known, motivating reconstructions at early time points (Lantz et al., 2003; Mălîia et al., 2016; Aydin et al., 2017; Plummer et al., 2019). Our results support the main findings of the multi-focal epilepsy case study of (Aydin et al., 2017), where zoomed MR imaging was guided by EMEG source analysis at the spike onset to reveal a fairly subtle focal cortical dysplasia in left fronto-central region, shown to be the trigger of the seizures. One difference is that in (Aydin et al., 2017) the single modality EEG or MEG failed, only EMEG was successful. Therefore, the use of complementarity can be of utmost importance particularly in low signal-to-noise ratio (SNR) situations as at the spike onset. In the present study, single modality EEG or MEG also localized into the epileptogenic zone but showed only sub-aspects of the overall source patch. Although it could not be shown experimentally here, we assume that the EMEG centroid at spike onset represents well the target position and orientation for individually optimized mc-tDCS, at least this hypothesis is supported by the highly significant reduction of IEDs (Figure 8).

### Limitations and outlook

We presented here a *proof-of-concept N-of-1* trial in a patient with focal epilepsy with the goals of (i) noninvasive reconstruction of the epileptogenic zone from EMEG data, (ii) noninvasive reduction of epileptic activity by EMEG-targeted and D-CMI optimized mc-tDCS, and (iii) parameterization of our processing pipelines for a future group study on focal epilepsy patients. Since all three goals were successfully achieved in the present case study, the group clinical trial using the parameters defined here is our most important future goal.

We must be aware that EMEG IEDs reconstruct the irritative zone, which does not necessarily have an overlap with the actual epileptogenic zone we are looking for and IED suppression does not imply seizure suppression. However, source reconstruction can of course also be applied to ictal data (Rullmann et al., 2009; Plummer et al., 2019). Furthermore, as (Karoly et al., 2016) hypothesized, spikes and seizures share one or more common regulatory mechanisms, as evidenced by their similar circadian patterns. In addition, in a large series of 1000 cases, (Rampp et al., 2019) showed that MEG IED reconstruction provides non-redundant information, which significantly contributes to long-term seizure freedom after epilepsy surgery. As the present study and other studies involving focal epilepsies due to cortical malformations such as focal cortical dysplasia (FCD) show (Aydin et al., 2017; Neugebauer et al., 2022), there is a substantial likelihood of overlap between both the irritative and the epileptogenic zones. As an outlook, the planned group clinical trial will show to what extent our proposed noninvasive approach can contribute to the reduction of ictal activity. This could not be clearly demonstrated in our case study, as under continued medication the seizure frequency was too low to measure success. The patient mentioned only one seizure, but it was of reduced severity. It should be mentioned here that (Kaufmann et al., 2021) reported a significant reduction in both interictal and ictal activity, but with reduced or discontinued medication, while recent studies of (Yang et al., 2020; Holmes et al., 2019) and also our study continued medication during tDCS.

With regard to long-term tDCS stimulation effects, (Auvichayapat et al., 2013) reported that a single session was associated with significant reductions in epileptic activity 48 h after stimulation and even four weeks later a statistically significant decrease in seizure frequency was detected. In (Kaufmann et al., 2021), tDCS induced a statistically significant reduction in IED and seizure frequency, which was outlasting 24 hours. (Zoghi et al., 2016a) studied temporal lobe epilepsy and found a statistically significant seizure frequency reduction even one month after stimulation. An even 4-month period of seizure reduction was found by (Zoghi et al., 2016b). (Yang et al., 2020) showed that the seizure reducing effect can be magnified and consolidated by application of 2 times 20 min daily stimulation (as also used here), which was superior to 20-min daily stimulation. Repeated applications of tDCS might thus have accumulative inhibitory effects on the epileptic network (Kaufmann et al., 2021). In our experiment, as shown in Figure 8, baseline IED frequency varied greatly during the 5 days of real stimulation and only slightly during the 5 days of sham stimulation. During real stimulation, the reductions in IED frequencies by mc-tDCS mainly added up on the first and last two days, whereas only after the one seizure with reduced severity on the evening of the second day, the baseline IED frequency on the third day was considerably higher, even more than twice the baseline frequency of the first day. In summary, the reduction in IED frequency added up except for the third day. We thus suspect that, without the seizure on day 2, the IED reductions would have added up so that a long-term stimulation effect could have been detected.

## 5. Conclusion

In this proof-of-principle N-of-1 study, a double-blind, sham-controlled stimulation experiment was performed in a patient with refractory focal epilepsy in the left frontal lobe close to Broca’s area. We applied, for the first time, combined EEG/MEG-targeted and distributed constrained maximum intensity (D-CMI) optimized multi-channel tDCS (mc-tDCS) in focal epilepsy and demonstrated that this non-invasive procedure can significantly reduce epileptic activity. Targeting was done in terms of both target location and orientation using combined EEG and MEG source analysis, as complementary information is contained in both modalities due to their different sensitivity profiles. For field modeling, we used the finite element method in a highly realistic anisotropic six-compartment head model calibrated for individual skull-conductivity and taking into account cranial burr holes from a previous invasive EEG. To the best of our knowledge, such a level of personalization in targeting, optimization and head modeling for noninvasive diagnosis and therapy of focal epilepsy has not yet been proposed and used before. Based on its success, our case study parameterizes a follow-up clinical trial for EMEG targeted and D-CMI optimized mc-tDCS in focal epilepsy.

## Data Availability

Fully anonymized data that support the findings of this study are available on request by the first author. The original data are not publicly available due to privacy and ethical restrictions.

## CRediT authorship contribution statement

**Marios Antonakakis**: Data curation, Software, Methodology, Formal analysis, Investigation, Visualization, Validation, Writing - original draft. **Fabian Kaiser**: EEG/mc-tDCS/EEG Data curation, Organization of blinding, Statistics, Investigation, Validation, Writing – review & editing. **Stefan Rampp**: Conceptualization, Funding acquisition, Ethics, Supervision, Spike marking, Writing – review & editing. **Stjepana Kovac**: Ethics, Spike marking, Writing – review & editing. **Heinz Wiendl**: Resources. **Walter Stummer**: Surgery, Resources. **Joachim Groß**: Resources, Writing – review & editing. **Christoph Kellinghaus**: Conceptualization, Ethics, Spike marking, Writing – review & editing. **Maryam Khaleghi-Ghadiri**: Ethics, Surgery, Writing – review & editing. **Gabriel Möddel**: Conceptualization, Funding acquisition, Ethics, Spike marking, Investigation, Validation, Writing – review & editing. **Carsten H. Wolters**: Conceptualization, Funding acquisition, Project administration, Ethics, Supervision, Validation, Software, Resources, Methodology, Writing - original draft.

## Acknowledgments

This work was supported by the Bundesministerium für Gesundheit (BMG) as project ZMI1-2521FSB006, under the frame of ERA PerMed as project ERAPERMED2020-227 PerEpi, by the German Research Foundation (DFG) through projects WO1425/7-1 and RA2062/1-1, by EU project ChildBrain (Marie Curie innovative training network, grant no. 641652), by DAAD project 57663920 and by the Onassis Scholarship Foundation. We acknowledge support from the Open Access Publication Fund of the University of Muenster. In addition, we thank Andreas Wollbrink and Christian Glatz for technical assistance and Luca-J. Bombardelli, Hildegard Deitermann, Juliana Gericks, Gabriele Kemper, Ute Trompeter, Pia Wenge and Karin Wilken for their help with the EEG/MEG/MRI and tDCS data collection.

## Footnotes

1 https://fsl.fmrib.ox.ac.uk/fsl/fslwiki/FSL

2 Seg3D: Volumetric Image Segmentation and Visualization. Scientific Computing and Imaging Institute (SCI): http://www.seg3d.org/

3 http://www.diffusiontools.com/documentation/hysco.html

4 http://vgrid.simbio.de/

5 SimBio: https://www.mrt.uni-jena.de/simbio/index.php/Main_Page and its integration into FieldTrip (see Vorwerk et al., 2018)

6 https://www.ibm.com/products/spss-statistics

